# Mask mandate and use efficacy for COVID-19 containment in US States

**DOI:** 10.1101/2021.05.18.21257385

**Authors:** Damian D. Guerra, Daniel J. Guerra

## Abstract

**Background:** COVID-19 pandemic mitigation requires evidence-based strategies. Because COVID-19 can spread via respired droplets, most US states mandated mask use in public settings. Randomized control trials have not clearly demonstrated mask efficacy against respiratory viruses, and observational studies conflict on whether mask use predicts lower infection rates. We hypothesized that statewide mask mandates and mask use were associated with lower COVID-19 case growth rates in the United States.

**Methods:** We calculated total COVID-19 case growth and mask use for the continental United States with data from the Centers for Disease Control and Prevention and Institute for Health Metrics and Evaluation. We estimated post-mask mandate case growth in non-mandate states using median issuance dates of neighboring states with mandates.

**Results:** Earlier mask mandates were not associated with lower total cases or lower maximum growth rates. Earlier mandates were weakly associated with lower minimum COVID-19 growth rates. Mask use predicted lower minimum but not lower maximum growth rates. Growth rates and total growth were comparable between US states in the first and last mask use quintiles during the Fall-Winter wave. These observations persisted for both natural logarithmic and fold growth models and when adjusting for differences in US state population density.

**Conclusions:** We did not observe association between mask mandates or use and reduced COVID-19 spread in US states. COVID-19 mitigation requires further research and use of existing efficacious strategies, most notably vaccination.

## Introduction

The COVID-19 pandemic has increased mortality and induced socioeconomic upheaval worldwide (1). Evidence-based containment strategies are warranted, given that age, obesity, cardiovascular disease, and diabetes are common comorbidities associated with severe COVID-19 symptoms (e.g., pneumonia, blood clots, cytokine storm), hospitalization, and death (2, 3). Respired droplets and aerosols containing SARS-CoV-2 are intuitive modes of community transmission (4). To reduce viral spread, governments have issued mandates to wear medical masks or cloth face coverings in public settings. From April to December 2020, 40 States of the United States issued mask mandates. Mask mandates have limited precedent, making efficacy unclear. Our first objective was to evaluate the efficacy of mask mandates in attenuating COVID-19 growth in US states.

Prior studies have conflicted on whether masks reduce COVID-19 spread. For USS Theodore Roosevelt crew, mask use was lower among COVID-19 cases compared with non-infected (56% vs. 81%) (5). There were no infections for 48% of universally masked patrons exposed to COVID-19 positive hair stylists (6), but PCR tests were not obtained for the other 52% of patrons (6), and first wave COVID-19 hospitalizations were no higher in public schools (high density with minimal masking) than elsewhere in Sweden (7). A randomized controlled trial (RCT) of Danish volunteers found no protective benefit of medical masks against COVID-19 infection (8). In RCTs before COVID-19, viral infections were not lower in Vietnamese clinicians who wore cloth or medical masks than in the control arm (9), and N-95 respirators (but not medical masks) protected Beijing clinicians from bacterial and viral diseases compared to no masks (10). Mask compliance in RCTs is not always clear (11). Mask use was 10% and 33% for Beijing households with and without intrahousehold COVID-19 case growth, respectively (12). This suggests greater mask use may reduce COVID-19 spread. Our second objective was to assess if mask use predicts lower COVID-19 case growth.

We assessed if mask mandates and compliance in US States predict statewide COVID-19 growth during the second and third infection waves (1 June 2020-1 March 2021). Controlling for infection wave timing with logarithmic and linear relative growth models, we found limited association between COVID-19 case growth and mask mandates or mask use before 1 October 2020, and no association during the subsequent and largest third wave. These findings do not support the hypothesis that statewide mandates and enhanced mask use slow COVID-19 spread. Pharmaceutical interventions (including recently available COVID-19 vaccines) provide alternative, evidence-based strategies to minimize COVID-19 related morbidity and mortality.

## Materials and methods

### Data Sources and Terms

We obtained total (confirmed and probable) COVID-19 cases up to 6 March 2021 for the 49 continental US states, normalized per 100,000 residents, from the Centers for Disease Control and Prevention (CDC) (13). To reduce reporting lag effects, we used 7-day simple moving means (e.g., the 7-day simple moving mean of cases on 31 March is the mean of daily cases between 28 March and 3 April). Hawaii was excluded because COVID-19 growth patterns deviated from those of continental US states. Confirmed and probable cases are defined by the Council of State and Territorial Epidemiologists. Confirmed cases require PCR amplification of SARS-CoV-2 RNA from patient specimens. Probable cases require one of the following: clinical and epidemiologic evidence, clinical or epidemiologic evidence supported by SARS-CoV-2 antigen detection in respiratory specimens, or vital records listing COVID-19 as contributing to death. Total PCR tests for each state were obtained from Worldometers on 25 May 2021 (14).

Mask mandates are statewide emergency executive public health orders requiring nose and mouth coverings in public settings in more than 50% of counties within a state (15, 16). We assigned US states to one of five quintiles based on when mandates went into effect (effective dates): 18 April-16 May 2020 (Q1), 29 May-3 July 2020 (Q2), 8 July-27 July 2020 (Q3), 1 Aug-9 Dec 2020 (Q4), or no statewide mandate as of 6 March 2021 (Q5). Effective dates were obtained from US state executive and health departments and press releases (available upon request).

We assessed mask use with the University of Washington Institute for Health Metrics and Evaluation (IHME) COVID-19 model site (17), which estimates daily compliance from Premise, the Facebook Global Symptom Survey (University of Maryland), the Kaiser Family Foundation, and the YouGov Behavior Tracker Survey. Mask use is the percentage of people who always wear masks in public settings. We assigned US states to mask quintiles based on the mean percent mask use from 1 Jun-1 Oct 2020 (Summer) or from 1 Oct 2020-1 Mar 2021 (Fall-Winter).

To assess geographic differences, we assigned each US state to one of five regions: Northeast (Connecticut, Delaware, Massachusetts, Maryland, Maine, New Hampshire, New Jersey, New York, Pennsylvania, Rhode Island, and Vermont); Midwest (Illinois, Indiana, Iowa, Kentucky, Kansas, Michigan, Minnesota, Missouri, Ohio, West Virginia, Wisconsin); Mountains-Plains (Colorado, Idaho, Montana, Nebraska, New Mexico, North Dakota, Oklahoma, South Dakota, Utah, Wyoming); South (Alabama, Arkansas, Florida, Georgia, Louisiana, Mississippi, North Carolina, South Carolina, Tennessee, Texas, Virginia); and Pacific (Alaska, Arizona, California, Nevada, Oregon, Washington).

### Growth Rate Calculation

COVID-19 growth has been modeled logarithmically (15, 18, 19) and linearly (19, 20). Therefore, we calculated COVID-19 case growth for each US state by measuring percent natural logarithmic (Ln Growth) and percent linear (Fold Growth) relative growth rates:

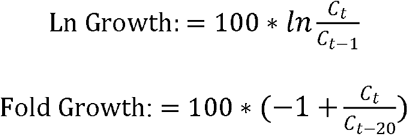

Where C_t_, C_t-1_, and C_t-20_ are total normalized cases on a day, the prior day, and 20 days prior, respectively. We determined adjusted population density by calculating the weighted mean of each state’s urban (U) and rural (R) population density using the following formulas:

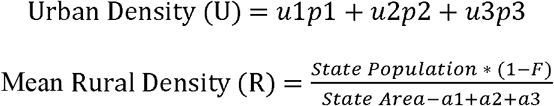

For the three most populous urban regions in each US state, we obtained urban population density (*u*; people/square mile in an urban area), urban land area (*a*; size of urban area in square miles), and population proportion (*p*; fraction of combined urban population) via 2010 US Census Bureau estimates (21). For some states, two rather than three urban regions were used. The proportion of urban (F) and rural (1-F) population of each state was similarly obtained (21). We thus calculated adjusted population density of each state as:

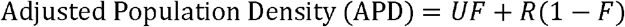

To assess association between population density and growth rates, we multiplied Fold Growth by the inverse of normalized APD:

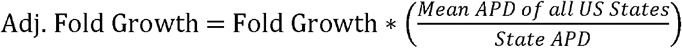

We defined minima and maxima (extrema) as the lowest and highest growth rates between the end of the Summer wave and the height of the Fall-Winter wave. Ln Growth extrema comprised 20-day windows when Ln Growth rates were lowest or highest:

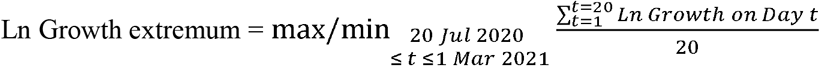

Fold Growth extrema similarly comprise the lowest and highest Fold Growth:

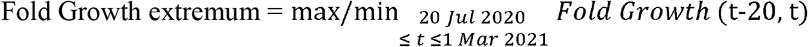

For each state, surges are differences between maxima and minima (relative growth rate increase), masks at extrema are the 20-day mean mask use at minima and maxima, and Δ Masks is the percent change in mask use between maxima and minima.

To assess association between mandates and growth rates in the 48 contiguous states (excluding Alaska and Hawaii), we determined Ln, Fold, and Adj. Fold Growth between 1 March 2021 (C_301_) and the mandate effective date (C_M_) for US states in mandate quintiles 1-3:

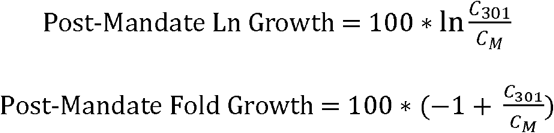

For states in quintiles 4-5, *modeled effective dates* are medians of actual dates among bordering states of mandate quintiles 1-3. For example, the modeled effective date of Tennessee (10 July) is the median of effective dates of Arkansas (20 July), Alabama (16 July), Kentucky (10 July), North Carolina (26 June), and Virginia (29 May).

For each state, Summer 2020 (1 June-1 Oct) and Fall-Winter 2020-21 (1 Oct-1 Mar) mask use is mean mask use between these dates. Cases on 1 June or 1 Oct were the 20-day mean total normalized cases on these two dates. We likewise defined Summer and Fall-Winter case growth using Ln and Fold Growth formulas:

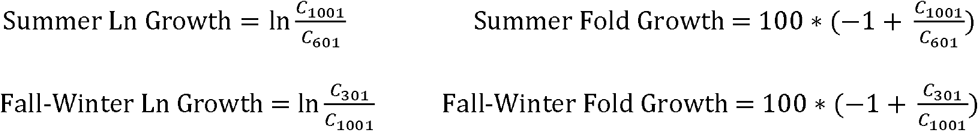

Where *c*_601_, *c*_1001_, and *c*_301_ are total normalized cases on 1 June 2020, 1 October 2020, and 1 March 2021, respectively.

### Statistics

We used Prism 9.2 (GraphPad; San Diego, CA) to construct figures and perform null hypothesis significance tests, for which the significance threshold was p < α = 0.05 (**Worksheet D in S1 Table**). Error bars denote standard deviations, 95% confidence intervals, or interquartile ranges as indicated in figure legends. We performed D’Agostino-Pearson tests to assess normality of residuals.

To evaluate mask mandate and use efficacy among categories (mandate effective date or mask use quintiles), we performed ordinary one-way ANOVA with Tukey posttests. For non-normal data, we performed Kruskal-Wallis with Dunn posttests. For two sample comparisons (e.g., **Fig. 3G, J**), we conducted two-tailed t tests or Mann-Whitney tests for normal and non-normal data, respectively. This decision tree conforms with recommended practices for datasets of N > 5 (22).

For interval variable associations, we performed ordinary least squares (OLS)-simple linear regression with null hypotheses of zero slope. Infectious disease research has employed OLS previously (23, 24), with linear and ln-linear models reported in recent COVID-19 studies (25, 26). For the Summer wave, Northeast states were excluded because they deviated from other states with respect to total cases and growth covariation. We used weighted least squares (WLS) for heteroscedastic data, as determined by the GraphPad Prism Test for Homoscedasticity. Regardless of statistical significance, R^2^ values denote coefficients of determination for lines of best fit with unconstrained slopes.

## Results

### COVID-19 growth rates vary with time

With the aim of reducing COVID-19 case growth, 40 US states enacted mask mandates in 2020. We wondered if mask mandate timing affected COVID-19 growth patterns. To identify patterns of COVID-19 growth, we graphed natural logarithmic (Ln) Growth of COVID-19 in US states as a function of time (**Worksheet A in S1 Table**). We observed six phases of COVID-19 growth up to 6 March 2021: first wave (before May 2020), Spring minimum (May-June 2020), Summer wave maximum (June-August 2020), post-Summer minimum (August-October 2020), Fall-Winter wave maximum (October-January 2020), and third minimum (March 2021) (**Fig. 1A and S1 Fig**). Hawaii growth patterns deviated from those of continental US states and was thus excluded from further analysis. Regardless of mask mandate effective date quintile, Ln growth patterns were comparable for all continental US states, and there was no association between normalized total cases and PCR tests (**S1 Fig**).

**Fig 1.**
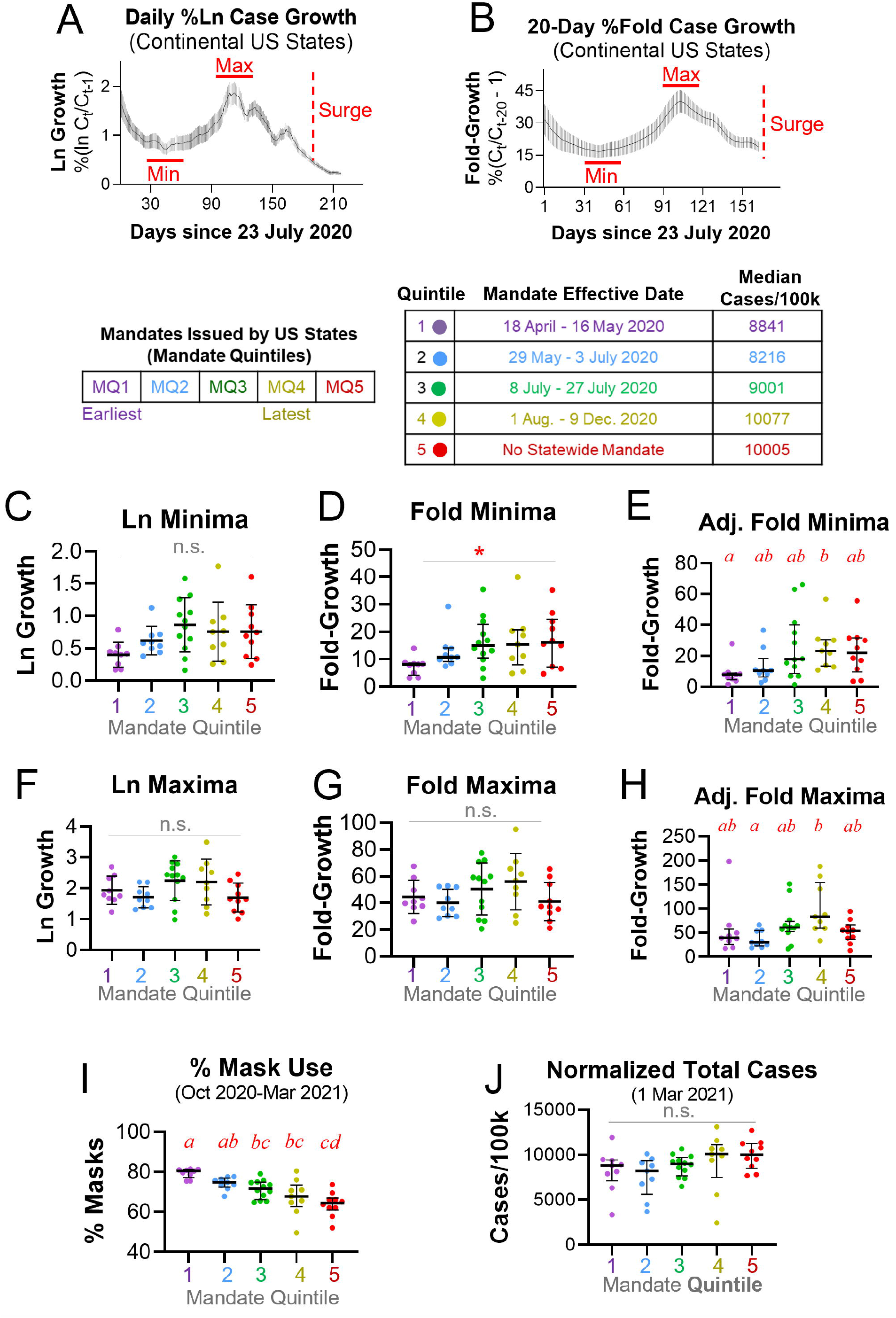
Earlier mask mandates are not consistently associated with lower COVID-19 growth rates in continental US States. A-B. Natural logarithmic (A) and Fold (B) COVID-19 growth in continental US states. Red horizontal lines denote growth rate minima (Min) and maxima (Max) between Summer and Fall-Winter waves. Surge: growth rate increase between Min and Max. C. Ln minima were not associated with the time quintile of a state’s mandate effective date (MQ). D. Fold minima trended lower in MQ1 than MQs 3 and 5. E. Adjusted Fold minima were lower in MQ1 than MQ4 and indistinguishable for all other pairwise comparisons. F-G. Ln (F) and Fold (G) maxima were not associated with mandate effective date time quintiles. H. Adjusted Fold maxima were lower in MQ2 than MQ4 and indistinguishable for all other pairwise comparisons. I. States with earlier mask mandates exhibited greater mask use between Oct. 2020 and March 2021. J. Cases per 100,000 by 1 March 2021 were not associated with mandate effective date time quintiles. Different letters denote p<0.05 by Tukey tests after one-way ANOVA (C, F, G) or all pairwise comparison Dunn tests after Kruskal-Wallis (D, E, H-J). *: p<0.05 by Kruskal-Wallis. n.s.: not significant. Error bars: 95% confidence intervals (A-B), standard deviations (C, F, G), and interquartile ranges (D, E, H-J).

### Earlier mask mandates are not consistently associated with COVID-19 growth rates in US states

A recent study reported time-enhanced negative association between mask mandates and Ln Growth of COVID-19 (15), but simple Fold-Growth (an alternative COVID-19 metric (19, 20)) may be preferred for post-exponential, linear pandemic spread. PCR testing for COVID-19 was limited before Summer 2020 (27). Thus, to determine if US states with earlier mask mandates exhibited less COVID-19 spread, we examined both Ln Growth and Fold Growth at the post-Summer wave minimum and the Fall-Winter wave maximum (**Fig 1A-B**)—periods of low and high transmission, respectively. We assigned US states to one of five quintiles (MQ1-5), with MQ1 including states with the earliest mandates, MQ4 the latest mandates, and MQ5 states without mandates. Ln minima (p=0.07; **Fig. 1C**) and Fold minima (p=0.047; **Fig. 1D**) trended lower for earlier mandates. Fold minima was 3-fold higher in MQ4 than MQ1 after adjusting for population density (p=0.04), but all other pairwise comparisons were not significant (**Fig. 1E**). This suggested that mask mandate duration was a weak predictor of lower minimum growth. Ln maxima (p=0.23; **Fig. 1F**) and Fold maxima (p=0.19; **Fig. 1G**) did not differ among quintiles. Adjusting for population density, Fold maxima were 2.8-fold higher in MQ4 than MQ2 (p=0.01), but MQ1, 3, and 5 were indistinguishable (**Fig. 1H**), suggesting mask mandate duration was not associated with lower maximum growth. Likewise, surges (growth rate increases from minima to maxima) were MQ-independent for Ln (p=0.08) and Fold (p=0.13) models, and only MQ2 and MQ4 exhibited significantly different Fold surges with population density adjustment (p=0.03; **S2 Fig**). Most MQ4 states exhibited lower initial and Summer 2020 infection waves than Q1-2 (**S1 Fig**), suggesting high MQ4 growth rates could be an artifact of lower total cases. While there was strong positive association between earlier mandates and Fall-Winter mask use (p<0.001; **Fig. 1I**), total cases on 1 March 2021 were MQ-independent (p<0.07; **Fig. 1J**). Direct MQ1 vs. MQ5 comparison by t test uncovered a small (1.2-fold) and non-significant (p=0.078) difference in total cases. Taken together, these findings suggest that US state mask mandates were not associated with slower spread of COVID-19.

### Early mask mandates do not predict lower post-mandate COVID-19 growth in contiguous US states

Most US states enacted mandates during infection waves, which confounds assessment of effectiveness. To assess association between mandate effective date and post-mandate case growth, we compared growth after actual MQ1-3 mandates with growth after modeled MQ4-5 mandates up to 1 March 2021. For a MQ4-5 state, the modeled date was the effective date median of contiguous, common-border MQ1-3 states (**Fig. 2A**). Post-mandate growth in total cases was MQ-independent by Ln (p=0.43), Fold (p=41), and population density-adjusted Fold (p=0.15) models (**Fig. 2B**). Direct MQ1 vs. MQ5 comparison by Mann-Whitney test uncovered a small (1.3-fold) and non-significant (p=0.86) difference in adjusted fold-growth. Overall, we did not obtain an association between mandates and lower COVID-19 growth.

**Fig 2.**
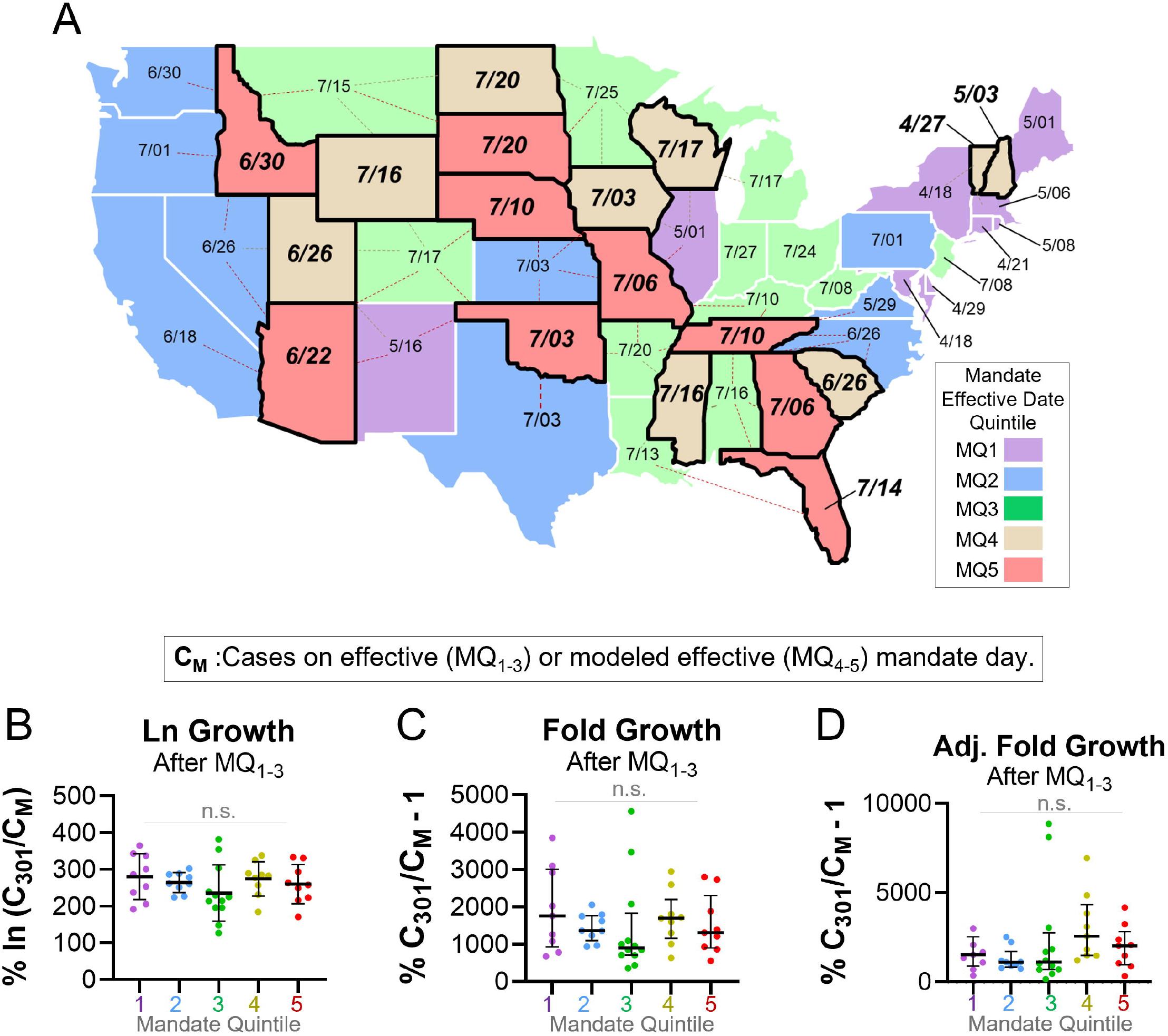
Earlier mask mandates are not associated with lower post-mandate COVID-19 growth rates in contiguous US states. A. Effective and modeled effective (***bold, italicized***) dates in 2020 for mask mandates in contiguous US states. State colors denote effective date time quintiles. Modeled dates of MQ4-5 states (late or no actual mandates) are medians of effective dates among bordering states of MQ1-3 (earlier mandates). Dashed lines denote MQ1-3 states that border a given MQ4-5 state. B-D. Between actual or modeled mandate effective dates and 1 March 2021, Ln Growth (B), Fold Growth (C), and population density-adjusted Fold Growth (D) were not associated with mandate effective date time quintiles. n.s.: not significant by one-way ANOVA (B) or Kruskal-Wallis (C-D). Error bars: standard deviations (B) and interquartile ranges (C-D).

**Fig 3.**
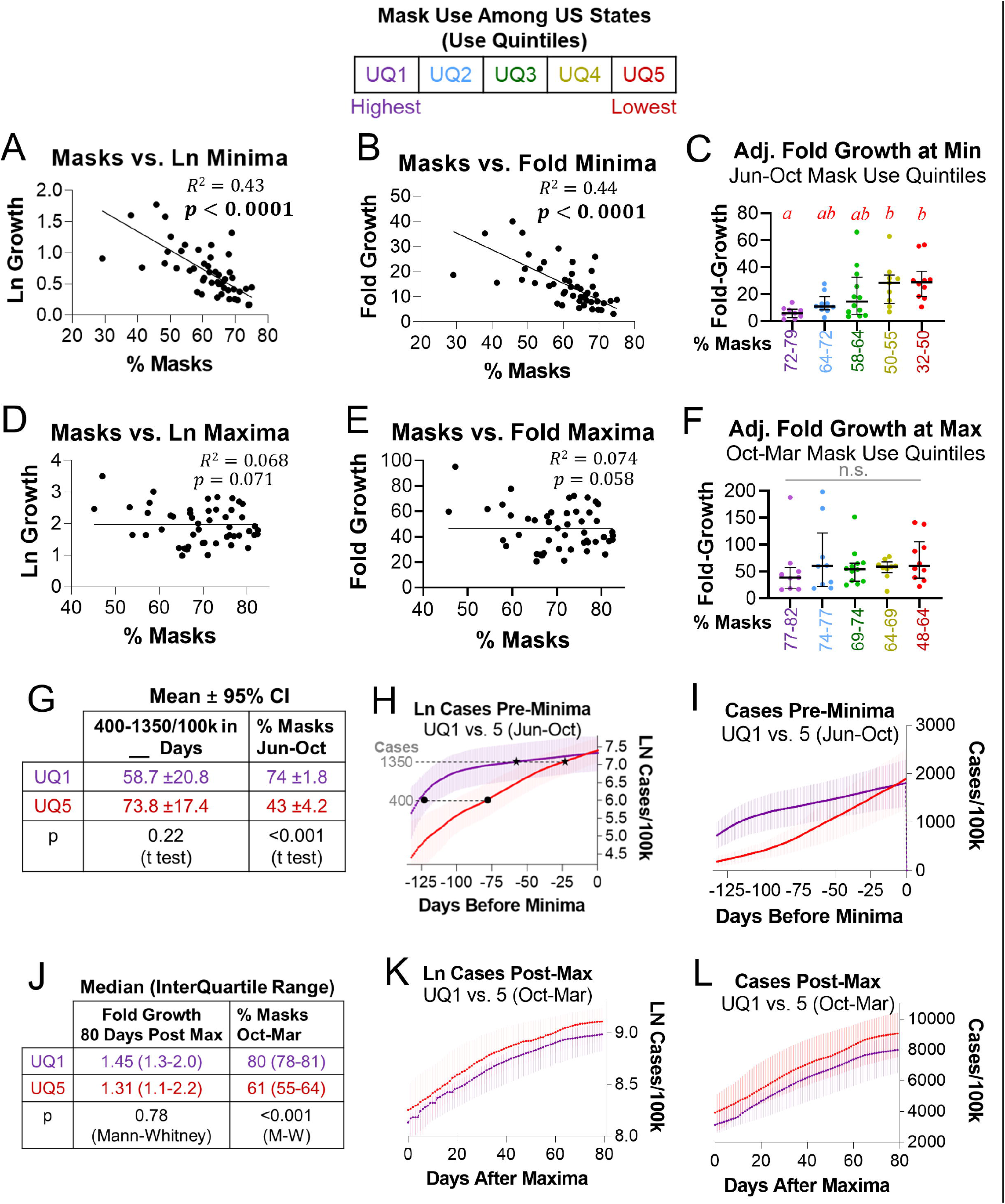
Mask use does not consistently predict COVID-19 case growth in continental US states. A-C. At minima, mask use was associated with lower ln (A), fold (B), and population density-adjusted fold (C) growth rates. D-F. At maxima, mask use was not associated with ln (D), fold (E), or population density-adjusted fold (F) growth rates. G. States in June-Oct. 2020 mask use quintiles (UQ) 1 and 5 grew from 400 to 1350 normalized cases at indistinguishable rates before minima. H-I. Ln cases (H) and cases (I) vs. time for UQ1 and UQ5. J. States in Oct. 2020-March 2021 mask use UQ1 and UQ5 exhibited indistinguishable Fold Growth 80 days after maxima. K-L. Ln cases (K) and cases (L) vs. time for UQ1 and UQ5. Simple linear regression used weighted (A-B) or ordinary (D-E) least squares. R^2^ values refer to unconstrained lines of best fit. Different letters denote p<0.05 by all pairwise comparison Dunn tests after Kruskal-Wallis (C, F). n.s.: not significant. Error bars: Interquartile ranges (C, F, J, K) and 95% confidence intervals (G-H).

### Mask use is not associated with most state COVID-19 case growth

We speculated that statewide mask use, rather than mask mandates per se, may predict lower COVID-19 growth rates. The Institute of Health Metrics and Evaluation (IHME) provides robust estimates for mask use (defined as the percentage of people who always wear masks in public settings) (17). By simple linear regression, mask use was associated with lower Ln, Fold, and adjusted Fold minima (p<0.0001; **Fig 3A-B** and **S3A Fig**). To better understand this trend, we assigned US states to one of five mask use quintiles (UQ1-5), with UQ1 including states with the highest mask use and UQ5 states with the lowest mask use. UQ5 exhibited a 3.4-fold greater adjusted Fold minimum than UQ1 (p=0.002; **Fig 3C**), suggesting potential association between mask use and COVID-19 spread at minima. By contrast, mask use was not associated with Ln (p=0.071), Fold (p=0.058), or adjusted Fold (p=0.076) maxima (**Fig 3D-E** and **S3D Fig**). Adjusted Fold maxima were also UQ-independent (p=0.56; **Fig 3F**), with direct UQ1 vs. UQ5 comparison by Mann-Whitney test uncovering a modest (1.5-fold) and non-significant (p=0.16) difference in maxima. This suggests that mask use is not associated with COVID-19 spread at maxima.

We wondered why mask use was associated with lower minimum but not lower maximum growth rates. Mask use was not associated with total cases at Ln minima (p=0.54) or maxima (p=0.086; **S3C-D Fig**), indicating potential confounders in the mask-minimum growth relationship. Excluding Northeast states, which exhibited the largest first waves and July 2020 seroprevalence (13, 28), total cases predicted lower Ln minima (p=0.001; **S3E Fig**). This suggested that the link between mask use and lower minima may be an artifact of the tendency for faster case growth to occur at lower case prevalence. In support of this, for 1 June – 1 Oct. 2020 mask use quintiles, normalized cases grew from 400 to 1350 at similar rates for UQ1 (which includes eight Northeast states) and UQ5 (p=0.22; **Fig. 3G**). UQ5 exhibited exponential growth and reached these case totals ∼50 days after UQ1 (**Fig 3H-I**), further implying that higher growth rates may reflect lower total cases in low mask use states before minima. 0-80 days after Ln maxima, when total case differences were smaller among states, UQ1 and UQ5 exhibited indistinguishable growth rates (p=0.78; **Fig 3J**; 1 Oct. 2020 – 1 March 2021 mask use quintiles). Growth was post-exponential for both UQ1 and UQ5 during this period (**Fig 3K-L**), and total cases predicted lower Ln maxima in all continental US states (p<0.0001; **S3F Fig**). Together, these data suggest that mask use is an unreliable predictor of COVID-19 growth in US states.

### Mask use does not predict Summer or Fall-Winter COVID-19 cumulative growth in US states

As expected, total cases were negatively associated with Ln growth in non-Northeast states for 1 June-1 Oct. 2020 and all continental US states for 1 Oct. 2020 – 1 March 2021 (p<0.0001; **S4A-B Fig**). We reasoned that even if mask use could not predict growth rate, mask use may be negatively associated with cumulative case growth. 1 June-1 Oct. 2020 (Summer) 1 Oct. 2020 – 1 March 2021 (Fall-Winter) represent two distinct COVID-19 growth waves (**Fig 4A-B**). Excluding Northeast states, masks were not associated with lower Summer growth using Ln (p=0.11; **Fig 4C**) or Fold (p=0.18; **S4C Fig**) models. Mask use trended with lower adjusted Fold Summer growth (p=0.05; **S4D Fig**). While adjusted Fold Summer growth was 3-fold higher in UQ4 than UQ1 (p=0.009), all other pairwise comparisons were not significant (**Fig. 4D**). Likewise, mask use was not associated with lower Fall-Winter growth using Ln (p=0.94; **Fig 4E**), Fold (p=0.91; **S4E Fig**), or adjusted Fold (p=0.71; **S4F Fig**) models, and adjusted Fold Fall-Winter growth was not significantly different among mask use quintiles (p=0.38; **Fig. 4F**). These data suggest that mask use is not consistently associated with Summer wave growth and not associated with Fall-Winter wave growth in US states. Furthermore, low Summer growth did not protect Northeast states from subsequent Fall-Winter growth. In summary, statewide SARS-CoV-2 transmission waves appear independent of reported mask use (17).

**Fig 4.**
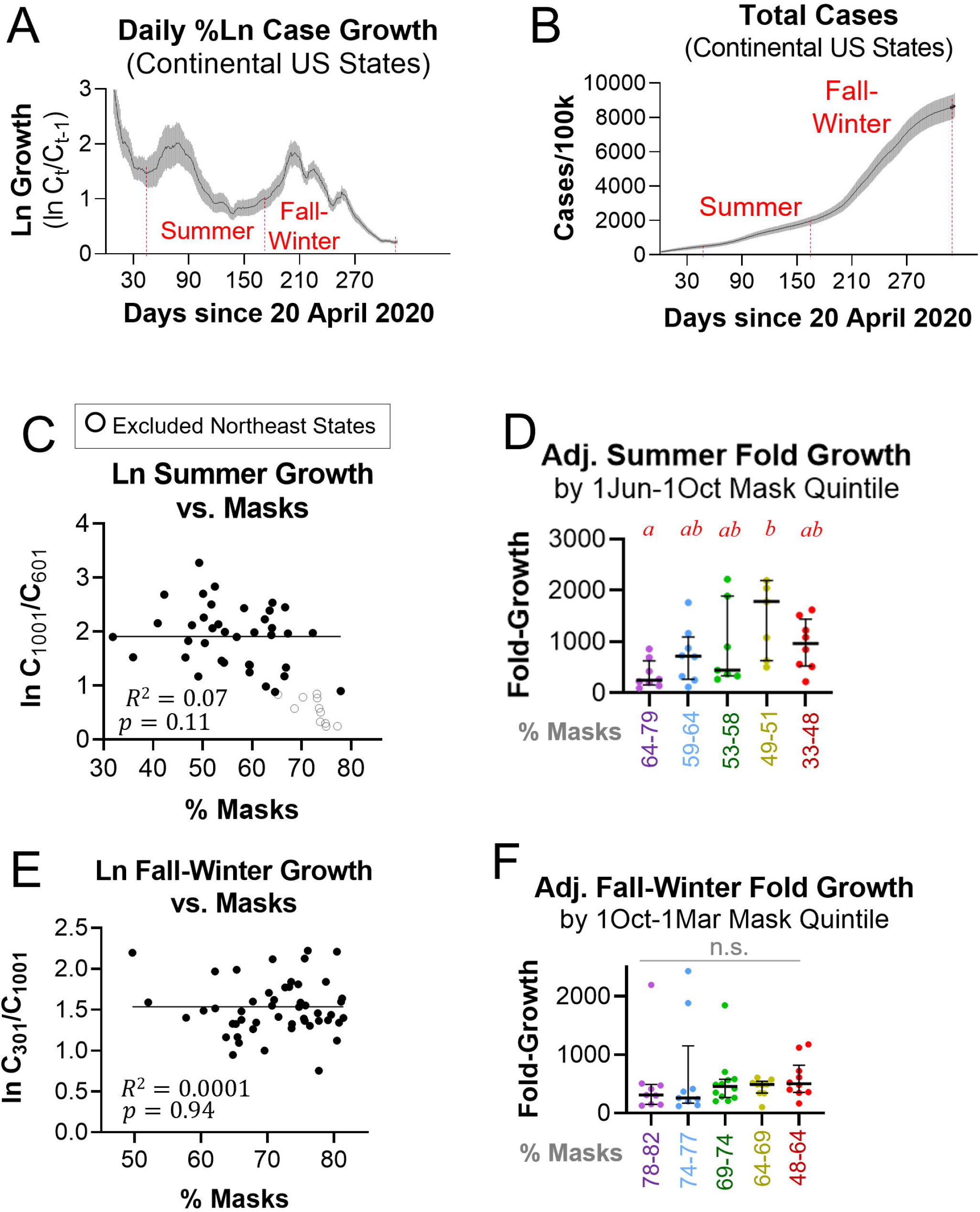
Mask use does not predict lower COVID-19 growth during the Summer or Fall-Winter waves. A-B. Ln Growth rate (A) and total COVID-19 cases (B) for continental US states from 20 April 2020 to 6 March 2021. Red vertical lines denote Summer (Jun-Oct 2020) and Fall-Winter (Oct 2020-Mar 2021) waves. C. Mask use does not predict Summer Ln Growth in non-Northeast states. D. In the Summer wave, population-adjusted Fold Growth was lower in Summer mask use UQ1 than UQ4 and indistinguishable among UQ2-5. E. Mask use does not predict Fall-Winter Ln Growth in continental US states. F. In the Fall-Winter wave, population-adjusted Fold Growth was indistinguishable among Fall-Winter mask use quintiles. Simple linear regression used ordinary least squares (C, E). R^2^ values refer to unconstrained lines of best fit. Different letters denote p<0.05 by all pairwise comparison Dunn tests after Kruskal-Wallis (D, F). n.s.: not significant. Error bars: 95% confidence intervals (A-B) and interquartile ranges (D, F). Solid circles (•): All continental US states. Hollow circles (○): Excluded Northeast states.

## Discussion

Our main finding is that mask mandates and use likely did not affect COVID-19 case growth. Mask mandates were associated with greater mask use but ultimately did not influence total normalized cases or post-mandate case growth. Higher mask use (rather than mandates per se) has been argued to decrease COVID-19 growth rates (11). While compliance varies by location and time, IHME estimates are derived from multiple sources and densely sampled. Even when accounting for population density, higher mask use was not associated with lower Ln or fold maximum growth rates or lower Fall-Winter case growth among continental US states. By contrast, mask use-growth rate association was highly significant at minima. This antinomy warrants consideration. Mask use did not predict normalized cases at growth minima or maxima, whereas there were more cases in the highest than the lowest mask use quintile before minima. Northeast states exhibited the highest seroprevalence by July 2020 (28) and comprised 80% of the highest mask use quintile, suggesting that mask use may be a lagging indicator of case growth. At maxima, when case prevalence was similar among states, COVID-19 growth rates were also similar for the highest and lowest mask use quintiles. Thus, initial association between masks and lower COVID-19 growth rates that dissipated during the Fall-Winter 2020-21 wave is likely an artifact of fewer normalized cases begetting faster growth in states with coincidental low mask use.

There is inferential but not demonstrable evidence that masks reduce SARS-CoV-2 transmission. Animal models (29), small case studies (6), and growth curves for mandate-only states (16) suggest that mask efficacy increases with mask use (11). However, we did not observe lower growth rates over a range of compliance at maximum Fall-Winter growth (45-83% between South Dakota and Massachusetts during maxima) (17) when growth rates were high. This complements a Danish RCT from 3 April to 2 June 2020, when growth rates were low, which found no association between mask use and lower COVID-19 rates either for all participants in the masked arm (47% strong compliance) or for strongly compliant participants only (8). While N-95 respirators offer some protection against respiratory viruses (10), there is limited evidence for cloth and medical masks. Higher self-reported mask use protected against SARS-CoV-1 in Beijing residents (30), but RCTs found no differences in PCR confirmed influenza among Hong Kong households assigned to hand hygiene with or without masks (mask use 31% and 49%, respectively) (31). Medical and cloth masks did not reduce viral respiratory infections among clinicians in Vietnam (9) or China (10), and rhinovirus transmission increased among universally masked Hong Kong students and teachers in 2020 compared with prior years (32). These findings are consistent with a 2020 CDC meta-analysis (33) and a 2020 Cochrane review update (34).

Our study has implications for respiratory virus mitigation. Public health measures should ethically promote behaviors that prevent communicable diseases. The sudden onset of COVID-19 compelled adoption of mask mandates before efficacy could be evaluated. Our findings do not support the hypothesis that greater public mask use decreases COVID-19 spread. As masks have been required in many settings, it is prudent to weigh potential benefits with harms. Masks may promote social cohesion during a pandemic (35), but risk compensation can also occur (36). By obscuring nonverbal communication, masks interfere with social learning in children (37). Likewise, masks can distort verbal speech and remove visual cues to the detriment of individuals with hearing loss; clear face-shields improve visual integration, but there is a corresponding loss of sound quality (38, 39). Prolonged mask use (>4 hours per day) promotes facial alkalinization and inadvertently encourages dehydration, which in turn can enhance barrier breakdown and bacterial infection risk (40). British clinicians have reported masks to increase headaches and sweating and decrease cognitive precision (41). Survey bias notwithstanding, these sequelae are associated with medical errors (42). Future research is necessary to assess risks of long-term daily mask use (34). As COVID-19 remains a public health threat, it is also appropriate to emphasize interventions with demonstrated efficacy against COVID-19, most notably vaccination (43) and vitamin D repletion (44).

In conclusion, we found mask mandates and use to be poor predictors of COVID-19 spread in US states. Strengths of our study include assessing COVID-19 association with both mandates and reported use; evaluating both Ln and Fold growth models; accounting for population density differences; and measuring case growth after modeled mandate effective dates in states with late or no mandates. Our study also has key limitations. We did not assess counties or localities, which may trend independently of state averages. While dense sampling promotes convergence, IHME masking estimates are subject to survey bias. We only assessed one biological quantity (confirmed and probable COVID-19), but the ongoing pandemic warrants assessment of other factors such as hospitalizations and mortality. Importantly, our study does not disprove the efficacy of all masks in limited and controlled circumstances, such as properly worn N95 respirators. A recent study found that at typical respiratory fluence rates, medical masks decrease airway deposition of 10-20µm SARS-CoV-2 particles but not 1-5µm SARS-CoV-2 aerosols (45). Aerosol expulsion increases with COVID-19 disease severity in non-human primates, as well as with age and BMI in humans without COVID-19 (46). Together with enhanced vaccination rates, aerosol treatment with improved ventilation and air purification could help reduce the size of COVID-19 outbreaks.

## Data Availability

Numeric data is contained within the manuscript as supplemental Excel tables. Raw data are available from the Centers for Disease Control and Prevention (link: https://data.cdc.gov/Case-Surveillance/United-States-COVID-19-Cases-and-Deaths-by-State-o/9mfq-cb36) or the Institute for Health Metrics and Evaluation (link: https://covid19.healthdata.org/). GraphPad Prism files are available upon request.

https://covid19.healthdata.org/

## Acknowledgments

The authors thank Brandy Jesernik, Ashley Tracey, Jay Bhattacharya, Scott Atlas, and Erik Fostvedt for manuscript input.

## Supporting Information Legends

**S1 Fig**. A. Natural logarithmic COVID-19 growth rates in continental US states by mask mandate time quintile. Y-axis values are differences between natural logarithms of total cases on a day and natural logarithms of total cases on the prior day. States are listed by time quintiles at which mask mandates became effective statewide. Thin black and wide gray denote mean and 95% confidence intervals, respectively. Red vertical lines denote dates of mask mandate issuance. Red horizontal lines indicate growth rate minima (phase 4) and maxima (phase 5) between the Summer and Fall-Winter waves. B. There is no association between cases and COVID-19 PCR tests among US states (case and test totals as of 25 May 2021).

**S2 Fig. Earlier mask mandates are not consistently associated with lower COVID-19 growth surges in continental US States**. Ln (A) and Fold (B) surges were not associated with the time quintile of a state’s mandate effective date. C. Adjusted Fold maxima were lower in MQ2 than MQ4 and indistinguishable for all other pairwise comparisons. Different letters denote p<0.05 by all pairwise comparison Dunn tests after Kruskal-Wallis (C). n.s.: not significant by one-way ANOVA. Error bars: standard deviations (A, B) and interquartile ranges (C).

**S3 Fig. Inverse association between normalized cases and COVID-19 growth rates in continental US states**. A. At minima, mask use predicts lower population density-adjusted Fold Growth. B. At maxima, mask use does not predict population density-adjusted Fold Growth. C. Normalized cases do not predict mask use at minima. D. Normalized cases do not predict mask use at maxima. E. At minima, greater total cases predict lower growth rates in non-Northeast continental US states. F. At maxima, greater total cases predict lower growth rates in continental US states at maxima. Black circles (•): All continental US states. Hollow blue circles 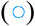: Excluded Northeast states. Solid blue circles 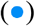: Included Northeast states. Red squares 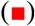: Midwest states. Green triangles 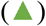: Mountain-Plains States. Grey triangles 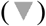: South states. Gold diamonds 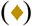: Pacific states. Simple linear regression used ordinary (A-D, F) or weighted (E) least squares and included all continental states except E (which excluded Northeast states). ε: Non-normal residuals by D’Agostino-Pearson test. R^2^ values refer to unconstrained lines of best fit.

**S4 Fig. Limited association between mask use and COVID-19 case growth during Summer and Fall-Winter waves**. A. Mask use does not predict Summer Fold Growth in non-Northeast states. B. Mask use is marginally associated with lower population-adjusted Fold Growth in non-Northeast states. C-D. Normalized cases were associated with lower Ln Growth during Summer (C) and Fall-Winter (D) waves in non-Northeast states (C) and all continental US states (D), respectively. E-F. Mask use does not predict Fall-Winter Fold Growth (E) or population-adjusted Fold Growth (F) in continental US states. Simple linear regression used ordinary least squares and excluded Northeast states (A, C, D) or included all continental US states (B, E, F). ε: Non-normal residuals by D’Agostino-Pearson test. R^2^ values refer to unconstrained lines of best fit.

**S1 Table. Supplemental Data Tables**. Worksheet A. 7-day rolling mean of total normalized cases (cases per 100,000 residents of each US state from CDC), 31 March 2020 - 1 March 2021. Worksheet B. Natural logarithmic daily case growth, 1 April 2020 – 1 March 2021. 20-day minima and maxima are denoted in red and gold, respectively. Mask mandate effective dates and modeled effective dates are provided for each state. Worksheet C. Population density calculations. See Materials and Methods for explanation of formulas. Worksheet D. 20-day Fold Growth, 12 August 2020 – 1 March 2021. Minima and maxima are denoted in red and gold, respectively. Population density-adjusted Fold Growth data are provided for each state. Worksheet E. Mask use for each US state at Ln and Fold extrema and during the Summer and Fall-Winter waves. Date range mask use values are simple arithmetic means of daily mask use over the specified date range. Numeric daily mask use (obtained from IHME) are provided.

