## Supplementary figures and images for "Mask mandate and use efficacy for COVID-19 containment in US States"

### Supplemental Figures 1-4

**S1 Fig**

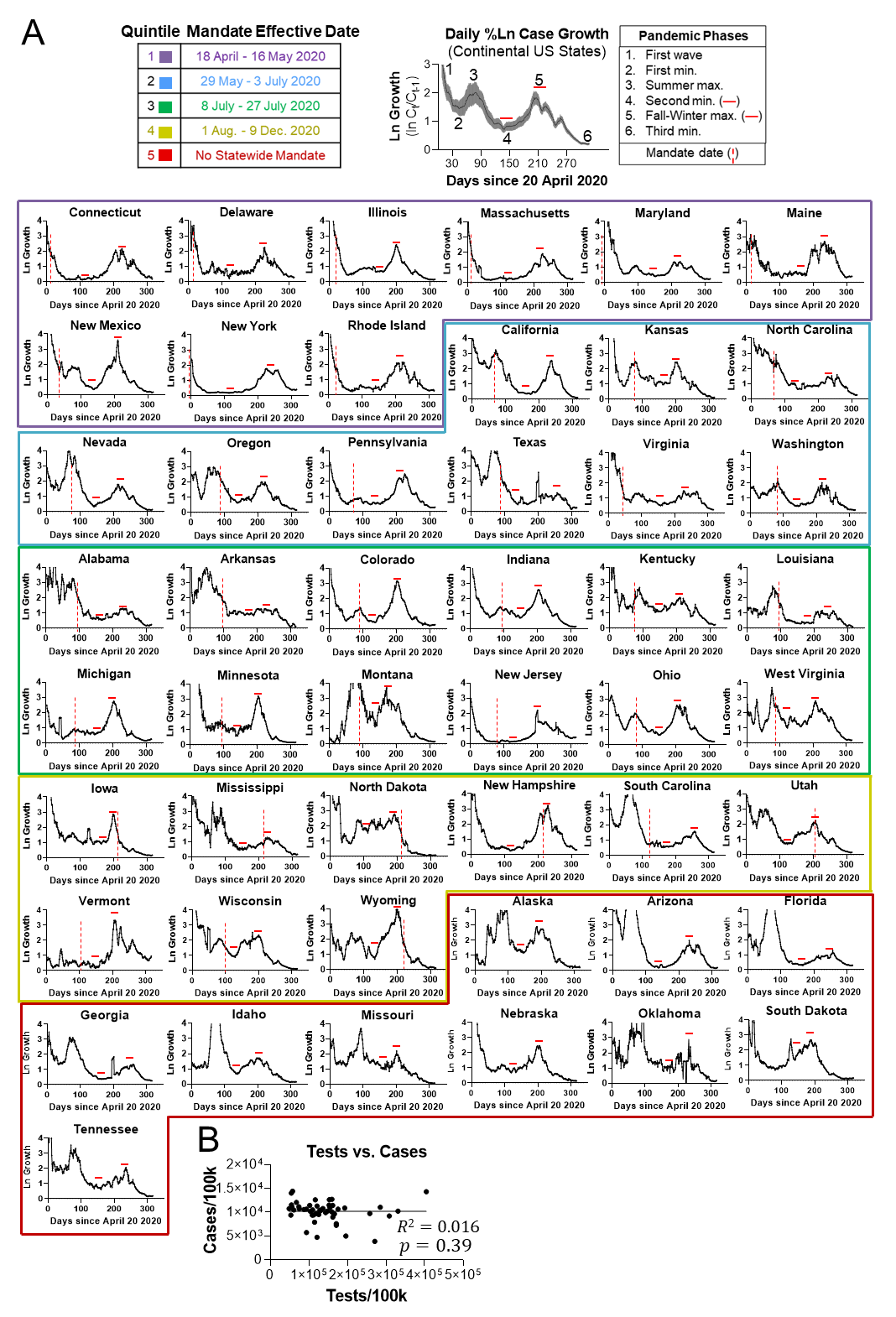

**S2 Fig**

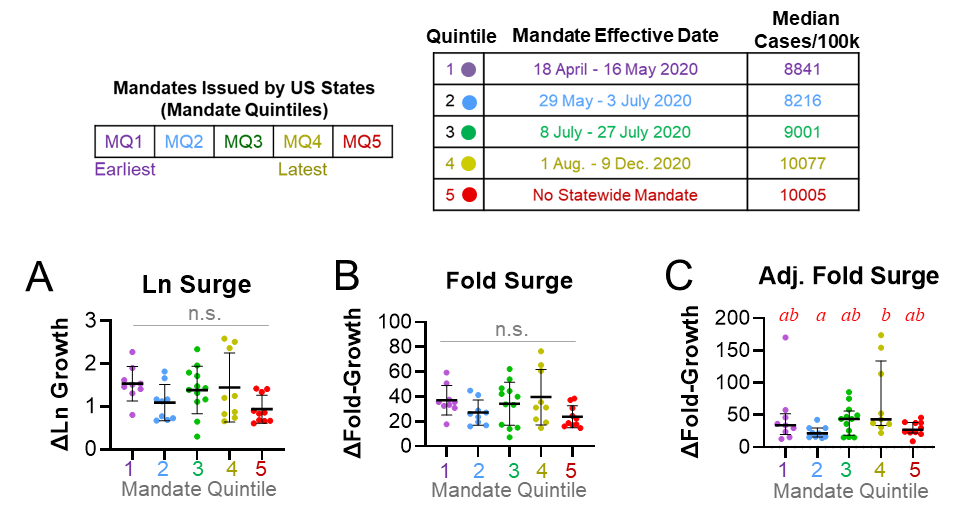

**S3 Fig**

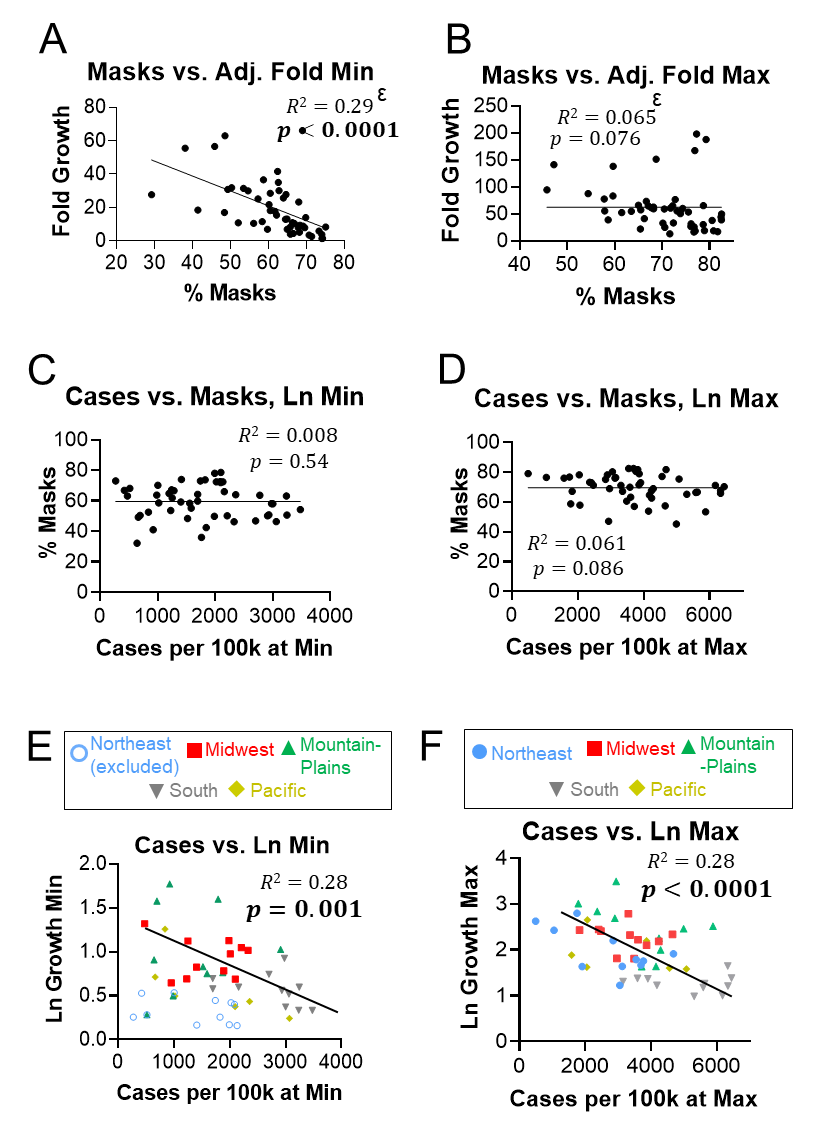

**S4 Fig**

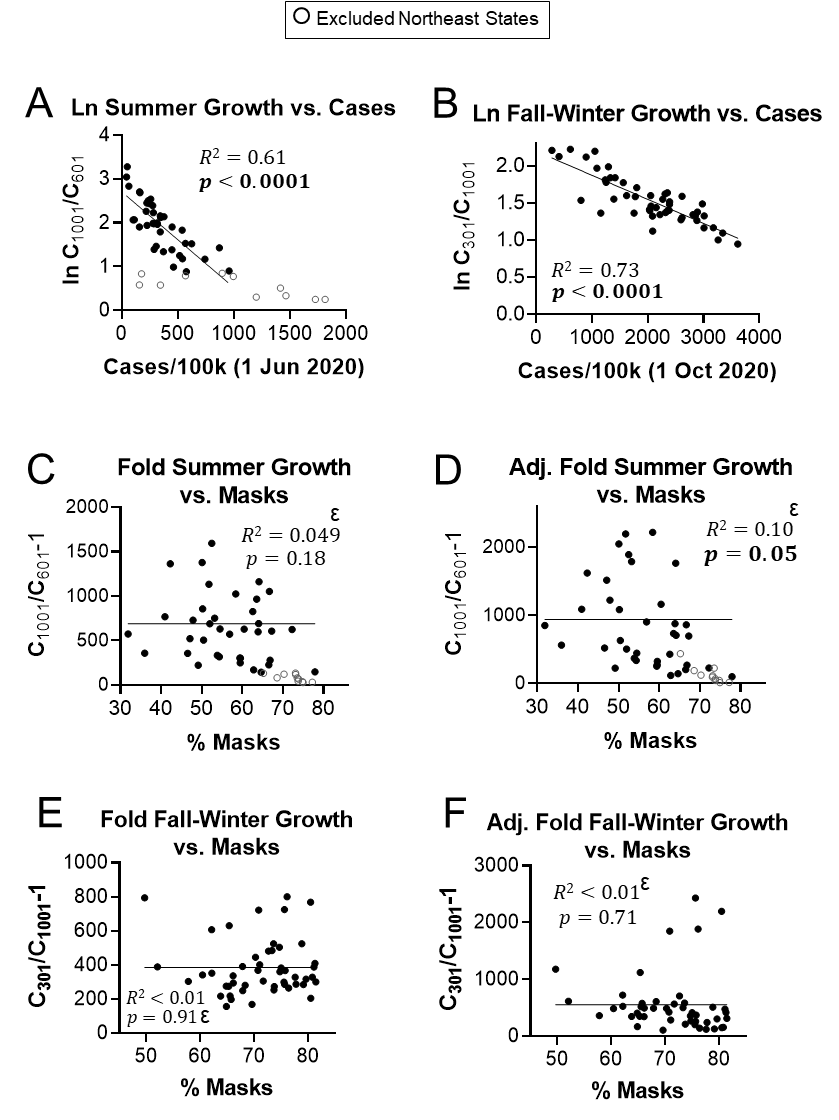
